# Triage assessment of cardiorespiratory risk status based on measurement of the anaerobic threshold, and estimation by activity limitation in patients with pulmonary arteriovenous malformations and hereditary haemorrhagic telangiectasia

**DOI:** 10.1101/2020.03.27.20045203

**Authors:** Saranya Thurairatnam, Filip Gawecki, Timothy Strangeways, Joseph Perks, Vatshalan Santhirapala, Jonathan Myers, Hannah C Tighe, Luke SGE Howard, Claire L. Shovlin

## Abstract

**Background:** Rapid triaging, as in the current COVID-19 pandemic, focuses on age and pre-existing medical conditions. In contrast, preoperative assessments use cardiopulmonary exercise testing (CPET) to categorise patients to higher and lower risk independent of diagnostic labels. Since CPET is not feasible in population-based settings, our aims included evaluation of a triage/screening tool for cardiorespiratory risk.

**Methods:** CPET-derived anaerobic thresholds were evaluated retrospectively in 26 patients with pulmonary arteriovenous malformations (AVMs) who represent a challenging group to risk-categorise. Pulmonary AVM-induced hypoxaemia secondary to intrapulmonary right-to-left shunts, anaemia from underlying hereditary haemorrhagic telangiectasia and metabolic equivalents derived from the 13-point Veterans Specific Activity Questionnaire (VSAQ) were evaluated as part of routine clinical care. Pre-planned analyses evaluated associations and modelling of the anaerobic threshold and patient-specific variables.

**Results:** In the 26 patients (aged 21-77, median 57 years), anaerobic threshold ranged from 7.6-24.5 (median 12.35) ml.min^-1^kg^-1^ and placed more than half of the patients (15, 57.7%) in the >11 ml.min^-1^kg^-1^ category suggested as “lower-risk” for intra-abdominal surgeries. Neither age nor baseline SpO_2_ predicted anaerobic threshold, or lower/higher risk categories, either alone or in multivariate analyses, despite baseline oxygen saturation (SpO_2_) ranging from 79 to 99 (median 92)%, haemoglobin from 108 to 183 (median 156)g.L^-1^. However, lower haemoglobin, and particularly, arterial oxygen content and oxygen pulse were associated with increased cardiorespiratory risk: Modelling a haemoglobin increase of 25g.L^-1^ placed a further 7/26 (26.9%) patients in a lower risk category. For patients completing the VSAQ, derived metabolic equivalents were strongly associated with anaerobic threshold enabling risk evaluations through a simple questionnaire.

**Conclusions:** Baseline exercise tolerance may override age and diagnostic labels in triage settings. These data support approaches to risk reduction by aerobic conditioning and attention to anaemia. The VSAQ is suggested as a rapid screening tool for cardiorespiratory risk assessment to implement during triage/screening.

**Key Messages:** 

**What is already known:**

- Alongside age, pre-existing medical conditions are perceived negatively during triage assessments, particularly if rare, and/or theoretically expected to influence cardiorespiratory risk;
- Anaesthetists use cardiopulmonary exercise testing to categorise patients to higher and lower risk independently to diagnostic labels, but this is not feasible in acute settings;
- Pulmonary arteriovenous malformations are an exemplar of a condition where, due to expected or measured abnormalities (hypoxaemia-low PaO_2_ SpO_2_), poor physiological capacity might be predicted.

**What this study adds:**

- Neither age nor usual SpO_2_ predicted lower/higher risk categories by anaerobic threshold, but haemoglobin-dependent indices of oxygen delivery to the tissues were associated with higher risk, offering opportunities for improvement by attention to anaemia and aerobic conditioning;
- Baseline exercise tolerance may override age and diagnostic labels in triage settings: the 13-point VSAQ Veterans Specific Activity Questionnaire (VSAQ) is suggested as a rapid screening tool for cardiorespiratory risk assessment.

## Introduction

Difficult triage decisions need to be made in many clinical settings involving large numbers of critically ill patients, as during the current COVID-19 pandemic. Such decisions are based on factors such as age and pre-existing medical conditions, in addition to acute observations and measurements. With the exception of certain common disease states, there is little evidence regarding associations with specific infections or complication risks. Diagnostic labels are generally linked under a single, negative umbrella of “pre-existing medical conditions”. There is particular concern that as for health insurance, lack of familiarity with rare diseases may lead to an inappropriately negative weighting, with no time to redress in an acute triage setting.

In pre-operative assessments, anaesthetists increasingly use cardiopulmonary exercise testing (CPET) to identify patients who may be unable to appropriately respond to increased cardiorespiratory demands of surgery due to reduced cardiorespiratory reserve.[1,2] The CPET-derived measure of anaerobic threshold (AT) of <11 ml.min^-1^ kg^-1^ has been identified in multiple studies and systematic reviews to be associated with adverse outcomes, mortality, and longer lengths of stay in a variety of surgeries, including intra-abdominal and intra-thoracic procedures.[2-4] The AT represents the point where ATP generation cannot be met by mitochondrial metabolism. It is considered a good measure as it reflects oxygen delivery and patient conditioning, and is not dependent on the patient’s motivation during exercise.[2]

The rapid assessment tool selected for evaluation was the Veteran’s Specific Activity Questionnaire (VSAQ [5]). This is a simple 13-point scale of activities of increasing difficulty whereby the user indicates which activity normally causes them to stop when performed for a period of time.[5] The activities correspond to metabolic equivalents (METs), and numerous studies show a good correlation with AT derived from cardiopulmonary exercise testing [6-8], mortality [9] and postoperative complications [10].

We focussed on one particular rare disease that provides an instructive example of potentially over-called cardiorespiratory risk. Pulmonary arteriovenous malformations (AVMs) are abnormal vascular connections between pulmonary arteries and veins, resulting in an anatomic right-to-left shunt.[11] Patients with pulmonary AVMs can demonstrate pronounced physiological abnormalities, including significant hypoxaemia, [12-18] increased minute ventilation 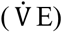 for given increases in CO_2_ production (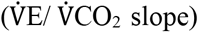) [16,17], high cardiac output states [19], and often iron deficiency and anaemia due to inadequate replacement of haemorrhagic iron losses from underlying hereditary haemorrhagic telangiectasia (HHT) [20,21] There is no published guidance on management of individuals with pulmonary AVMs or HHT undergoing anaesthesia, and each year, our service receives requests regarding suitability for surgery and insurance, and/or reports that surgery or insurance has been withheld because of the perceived risks of pulmonary AVMs/HHT.

Our goal was to evaluate commonly used assessment criteria and examine the potential role for a rapid assessment tool that could distinguish lower risk individuals in an emergency setting, based on usual cardiorespiratory status. The detailed study aims were to explore which variables may be associated with cardiorespiratory risk defined by the anaerobic threshold in order to inform triage and develop approaches to help guide pre-exposure [23] or pre-operative [1-4,10] management. Having recently applied the VSAQ to observational studies in patients with pulmonary AVMs and HHT, [17,18] we hypothesised that this could prove to be a useful risk categorising tool for triage purposes across wider patient groups.

## Methods

### General patient evaluations

With ethical approvals (Hammersmith and Queen Charlotte’s and Chelsea Research Ethics Committee (LREC 2000/5792), patient indices derived as part of the clinical assessment process in a pulmonary

AVM service at a single centre were examined as described elsewhere [14,15,17,18]. These included arterial oxygen content (CaO_2_) derived from SpO_2_ values measured in the erect posture breathing room air using the established formula:

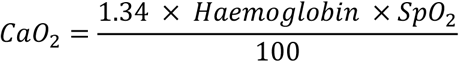

### Cardiopulmonary exercise tests

Previously reported cardiopulmonary exercise tests in patients with pulmonary AVMs where there had been striking variability in anaerobic thresholds [16,17] were reanalysed with a focus on triage/pre-operative assessments methods. Ethical approval had been granted by the National Research Ethics Service (NRES) Committee South West London REC3: 11/H0803 and NRES London Riverside Committee:15/L0/0590. Written informed consent had been obtained from all participants. Full methodological details are provided in [16,17].

### Veteran’s Specific Activity Questionnaire (VSAQ)

The VSAQ was administered as part of routine clinical care to patients for independent completion, using a modified version as presented in ***Figure 1***. Patient-reported activity limitations in the 13-point scale were converted to metabolic equivalents (METs) in which 1 MET equals the consumption of 3.5 ml O_2_ per kilogram of body weight. Metabolic equivalents (METs) were calculated from the VSAQ as in the original protocol,[5] and subsequent validations [6-10], by the formula:

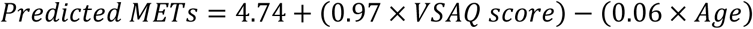

**Figure 1:**
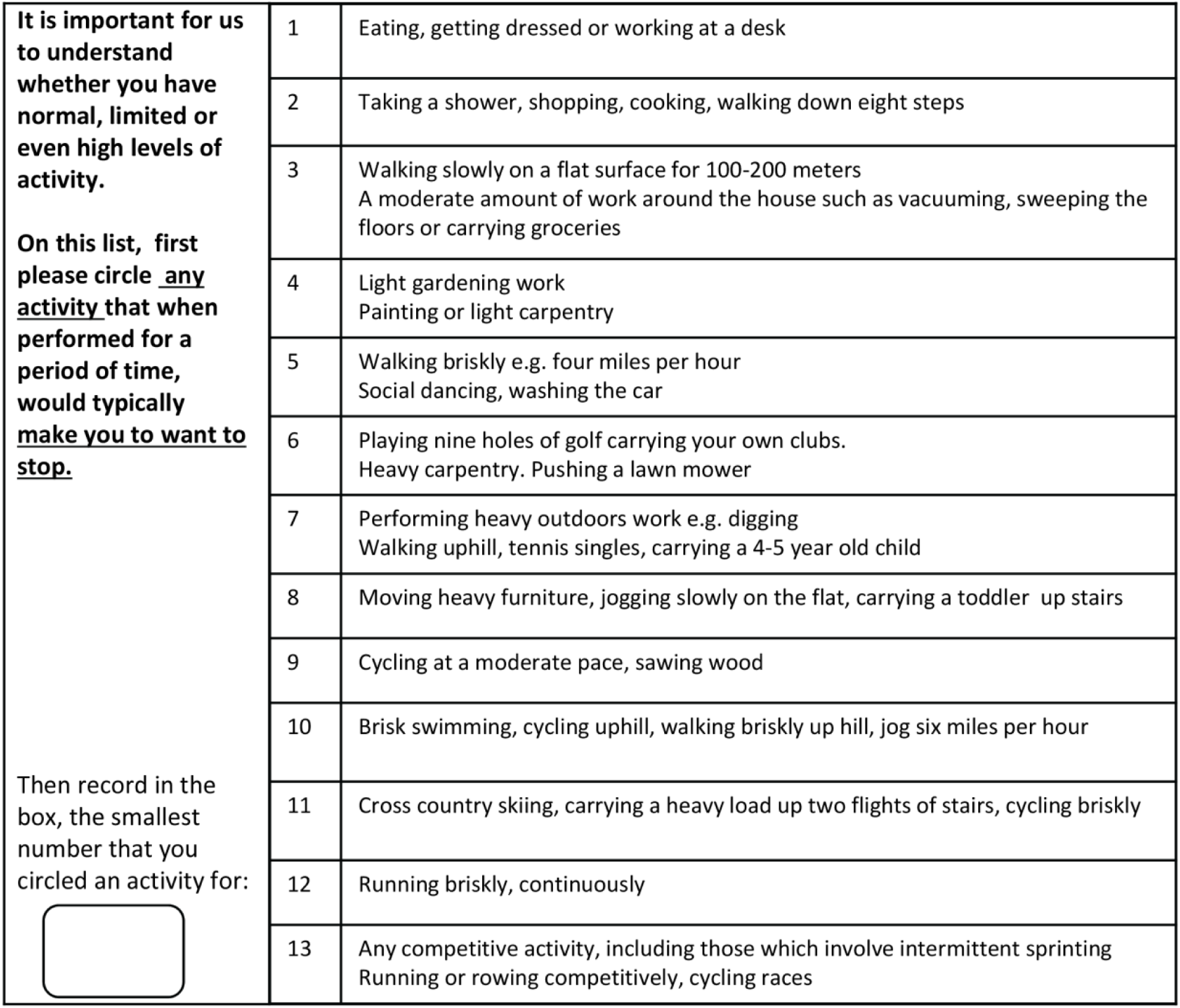
The UK-modified Veterans Specific Activity Questionnaire (VSAQ) *(adapted from [18] with authors’ permission)*

### Data Analysis

Statistical analyses were performed in Microsoft Excel and Stata IC versions 14 and 15 (Statacorp, Texas). Two-way analyses used Mann Whitney *U* test and three-way analyses Kruskal Wallis tests. Prior to data analyses, patients were pre-categorised based on the published anaerobic threshold delimiter of 11 ml.min^-1^ kg^-1^.[24] Since the risk categories may change in the future as more evidence becomes available, patients were also pre categorised above and below the median AT value.[24]. Additionally, to further support the robustness of data analysis, regression analyses were performed using A and log-transformed AT as the outcome variables (log-transformed AT had a more normal distribution, data not shown).

### Patient and Public Involvement statement

Patients were involved in earlier testing of the VSAQ [17,18] and aspects of design of the CPET protocols. Focussing of our data towards the triaging of patients was an outcome of inputs from British patients contacting us in March 2020, focussing on the question “Am I at High Risk?

## Results

### CPET Participant Demographics

The 26 patients with pulmonary AVMs comprised 16 male, 10 females, and were aged 21-77 (median 57) years. SpO_2_ ranged from 79 to 99 (median 92)%, haemoglobin from 108 to 183 (median 156)g.L^-1^, and body mass index (BMI) from 20 to 35.7 (median 26.1) kg.m^-2^. Comorbidities were present in 11 patients: three had known asthma or chronic obstructive pulmonary disease (COPD), one had sleep apnoea, and two had type 2 diabetes mellitus. In addition, three had suffered a previous stroke, transient ischaemic attack, or venous thromboemboli, one was in atrial fibrillation, one had well controlled hypertension, one was hypercholesterolaemic, one was significantly depressed, and two had benign prostatic hypertrophy.

### CPET Demographics identify a low risk group

As presented in ***Table 1***, based on the established anaerobic threshold delimiter of 11 ml.min^-1^ kg^-1^, more than half of the cohort with pulmonary AVMs (15/26, 57.7%) were categorised as pre-operative “low risk”, comprising 13 males and 2 females. The low risk group achieved a median 97% of their predicted maximum work rate compared to the high risk group median of 68% predicted (Table 1). Similarly, the median peak 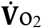 in the low risk group was 160% of the median in the high risk group.

The CPET-evaluated total oxygen consumption at peak exercise 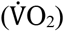, has also been used for high risk anaesthetic categorisation, noting that for reliable peak 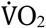 measurements, patients need to meet their point of maximal exercise. In the current study, all low risk patients identified by anaerobic threshold were also in a low risk category if defined by peak VO_2_< 20 ml.min^-1^.kg^-1^ [2] (data not shown).

### Age and usual SpO_2_ not associated with cardiorespiratory risk or anaerobic threshold

There was no difference in age between the low and high risk groups categorised by an anaerobic threshold delimiter of 11 ml.min^-1^ kg^-1^ (low risk mean 52.2 [95% confidence interval CI 43.8, 60.6] years, versus high risk mean 53.9 [95% CI 42.9, 64.9] years). ***Table 1*** and ***Figure 2A*** display the median values, the interquartile ranges (IQR), and 2 standard deviations. There was also no difference in age between the low and high risk groups categorised by upper/lower 50^th^ percentiles (lower risk mean 52.6 [95% CI 43.8, 61.5] years, versus higher risk mean 53.2 [95% CI 43.2, 63.3] years (***Table 2***)). In keeping with this, there was no detectable association between age and the absolute or log-transformed anaerobic threshold values (*p-*values >0.62, data not shown).

**Figure 2:**
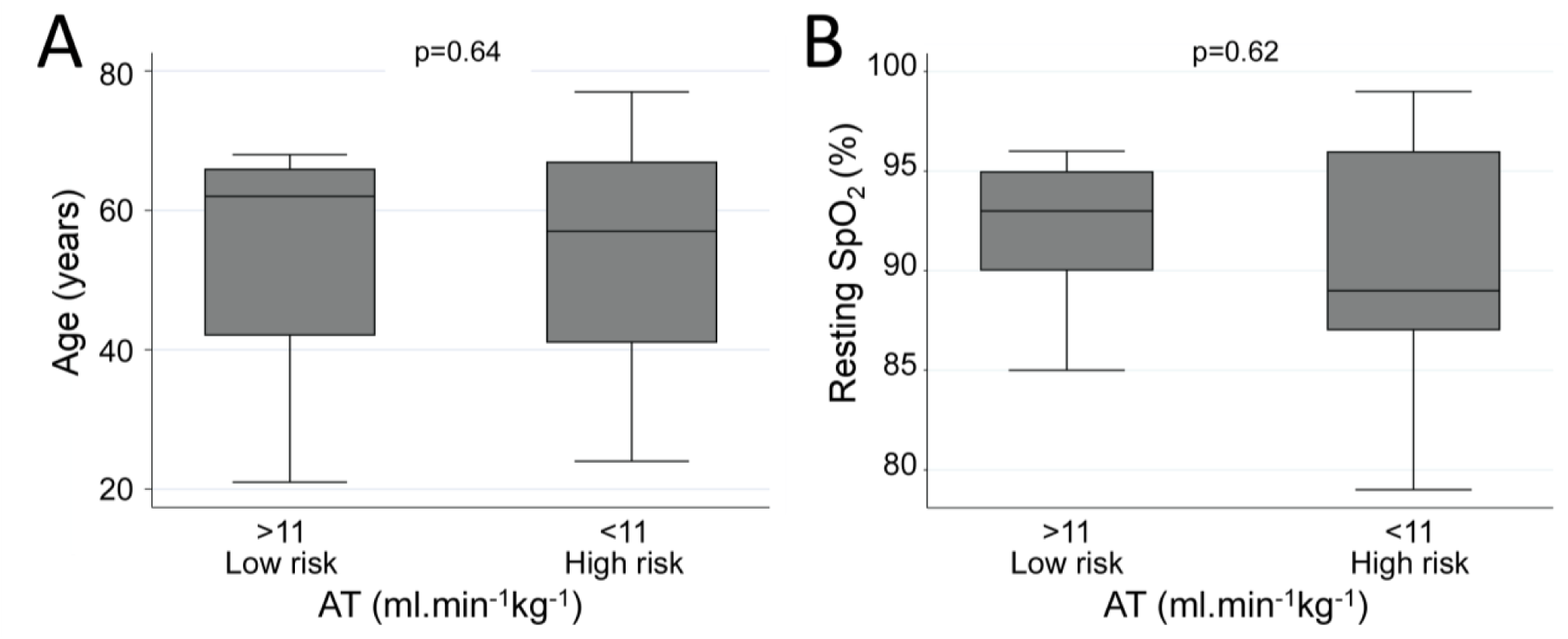
Age and oxygen saturation in low and high risk groups. *Box plots comparing values between the low risk and high risk anaerobic threshold (AT) groups for 26 pulmonary AVM patients, where high risk status was defined by AT lower than 11ml*.*min*^*-1*^*kg*^*-1*^: ***A)*** *Age (ys)*, ***B)*** *Resting SpO*_*2*_ *(%)*.*Boxes indicate the median and interquartile range (IQR), and error bars represent 2 standard deviations, with dots at the extremes representing outliers. P-values were calculated by Mann Whitney U test*.

Despite some very low resting SpO_2_ measurements, there was also no difference in pulse oximetry-measured oxygen saturation (SpO_2_) between the low and high risk groups: Categorised by an anaerobic threshold delimiter of 11 ml.min^-1^ kg^-1^, the means [95% CI] were 92 [90, 94]% for the low risk group, versus 91 [86, 95]% for the high risk group (***Table 1, Figure 2B***). Categorised by the upper/lower 50^th^ percentile groups, the respective means [95% CI] were 92.5 [90.3, 94.6]% for lower risk, versus 90.5 [87.1, 94.0]% for higher risk (***Table 2***). In crude and age-adjusted regression there was no detectable association between SpO_2_ and either category, or the absolute anaerobic threshold in ml.min^-1^ kg^-1^ or the log transformed values (*p-*values > 0.48, data not shown).

We concluded that assuming the study had sufficient power, neither age, nor more surprisingly SpO_2_ (in the setting of a compensated baseline state), in themselves, would be markers of a higher risk state in the cohort.

### Low haemoglobin, arterial oxygen content and oxygen pulse indicative of higher risk status

A different picture emerged when examining markers of oxygen delivery to the tissues, thus confirming that the study did have sufficient power to be discerning:

First, the higher risk groups had a trend towards lower haemoglobin, although there was some overlap in confidence intervals. Categorised by an anaerobic threshold delimiter of 11 ml.min^-1^ kg^-1^, the mean [95% CI] values were 159 [14.9, 17.0] g.L^-1^ for the low risk group compared to 144 [13.1, 15.6] g.L^-1^ for the high risk group (***Table 1, Figure 3***). By the upper/lower 50^th^ percentiles, the respective means [95% CI] were 158 [146, 171] g.L^-1^ for the lower risk group compared to 147 [135, 159] g.L^-1^ for the higher risk group (***Table 2***).

**Figure 3:**
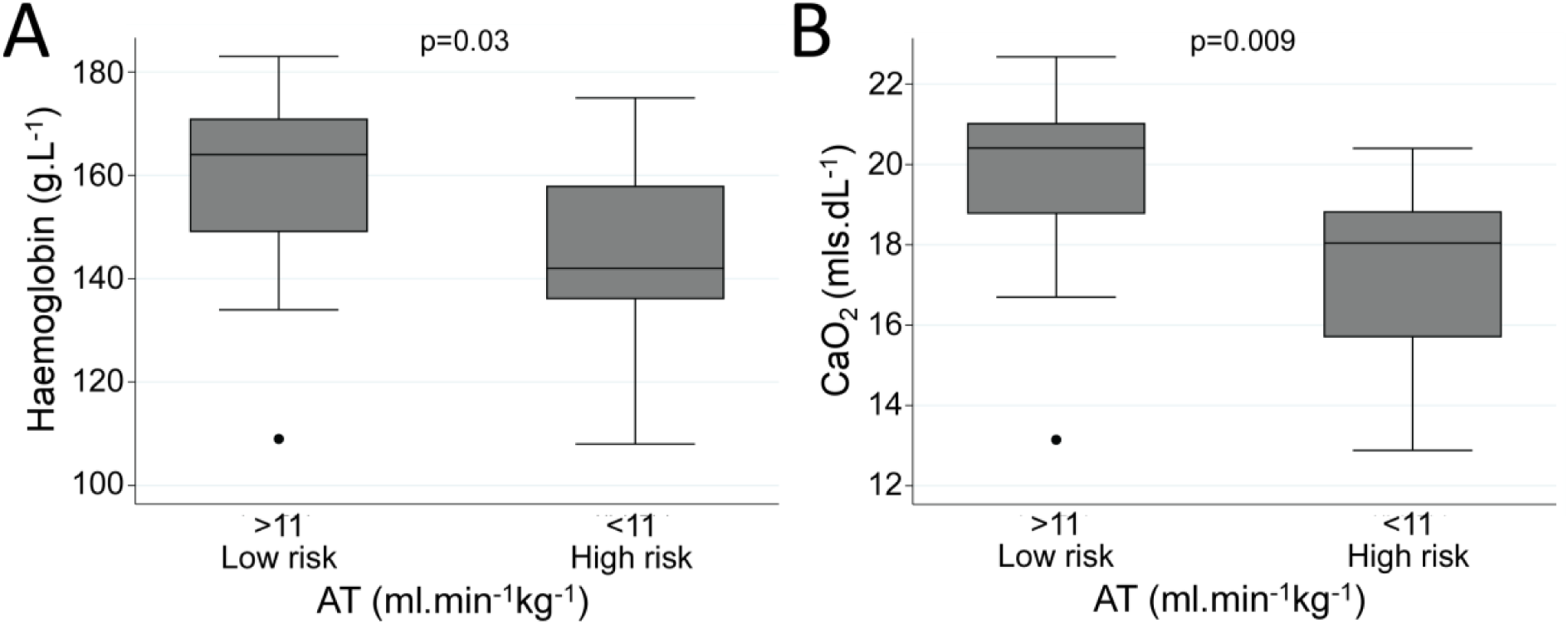
Haemoglobin and arterial oxygen content in low and high risk groups. *Box plots comparing values between the low risk and high risk anaerobic threshold (AT) groups for 26 pulmonary AVM patients, where high risk status was defined by AT lower than 11ml*.*min*^*-1*^*kg*^*-1*^: ***A)*** *Haemoglobin (g*.*dL*^*-1*^); ***B)*** *CaO*_*2*_ *(ml*.*dL*^*-1*^*). Boxes indicate the median and interquartile range (IQR), and error bars represent 2 standard deviations, with dots at the extremes representing outliers. P-values were calculated by Mann Whitney U test*.

More strikingly, the higher risk groups had significantly lower arterial oxygen content (CaO_2_) representing the oxygen content per unit volume of arterial blood, and calculated based on the oxygen carriage of 1.34mls per gram of fully saturated haemoglobin: Categorised by an anaerobic threshold delimiter of 11 ml.min^-1^.kg^-1^, the mean [95% CI] values were 19.6[18.4, 21.0] ml.dL^-1^ for the low risk group, but 17.4 [15.8, 19.0] ml.dL^-1^ for the high risk group (***Table 1, Figure 3***). Categorised by the upper/lower 50^th^ percentiles, the means [95% CI] were 19.6 [18.1, 21.0] ml.L^-1^ for the lower risk group but only 17.8 [16.4, 19.3] ml.L^-1^ for the higher risk group (***Table 2***).

Similarly, the higher risk group had a significantly lower mean oxygen pulse, representing the amount of oxygen extracted/ delivered per heart beat: Categorised by an anaerobic threshold delimiter of 11 ml.min^-1^ kg^-1^, the mean [95% CI] values were 14.4 [11.0, 17.3] ml.beat^-1^ for the low risk group, but 9.7 [6.7, 12.6] ml.beat^-1^ for the high risk group (***Table 1***). Categorised by the upper/lower 50^th^ percentiles, the mean [95% CI] values were 14.6[12.3, 16.9] ml.L^-1^ for the lower risk group but only 9.7 [7.2, 12.1] ml.L^-1^ for the higher risk group (***Table 2***).

Anaemia is very common, and readily correctable in clinical practice. We modelled whether increasing haemoglobin alone might allow patients to move from a high to lower risk category. In multivariate regression analysis, haemoglobin explained 57.3% of the variance in log-transformed AT (adjusted R^2^ 0.57). The regression coefficient of 0.053 (95% confidence interval 0.005, 0.100, p=0.031) implied that for each 1 g.dL^-1^ (10 g./L^-1^) rise in haemoglobin, the AT would rise by 0.76 ml.min^-1^.kg^-1^, and a haemoglobin rise of 2.5 g.dL^-1^ would increase AT by 1.9 ml.min^-1^.kg^-1^, moving 7 (63.6%) of patients from high to low risk. In the existing dataset, it was not possible to preselect all patients who would benefit using single resting demographics (data not shown).

Other measurements that were associated with the higher risk status were higher serum bicarbonate and higher minute ventilation 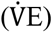 for given increases in CO_2_ production (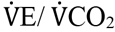 slope, Supplementary Figure 1). These are not currently amenable to therapeutic correction.

### VSAQ Score association with anaerobic threshold and risk categorisation

Having demonstrated that at least half of the patients with the rare pulmonary vascular abnormality would have their good exercise capacity and lower risk status readily identified were it feasible to perform CPET, we were conscious that in standard clinical practice, it is impractical to perform CPET on every patient. Usual activity could however be analysed by the VSAQ. As in data published for other general population cohorts,[5-9] for pulmonary AVM patients also completing the VSAQ, there was a good association between the previous CPET-derived AT and METs derived from the VSAQ (Supplementary Figure 3).

We used the derived relationship between and AT and VSAQ to model the expected cut off by age on the VSAQ that might indicate an individual in a lower risk category based on the established AT of 11 ml.min^-1^.kg^-1^. As noted in ***Figure 4***, this differs by patient age such that a VSAQ of 8 would be suggestive of lower risk irrespective of age, whereas older individuals in the “best” New York Heart Association (NYHA) category [25] could still fall into the higher risk category defined by AT <11ml.min^-1^.kg^-1^. In younger individuals, lower risk would be assigned even in the setting of more limited exercise capacity, (VSAQ 4-7, NYHA II). In other words, the VSAQ provided sufficient granularity to indicate where age-related physiology would be ’offset’ in particularly active older adults, and where there may be more concern for a much less active younger individual.

**Figure 4:**
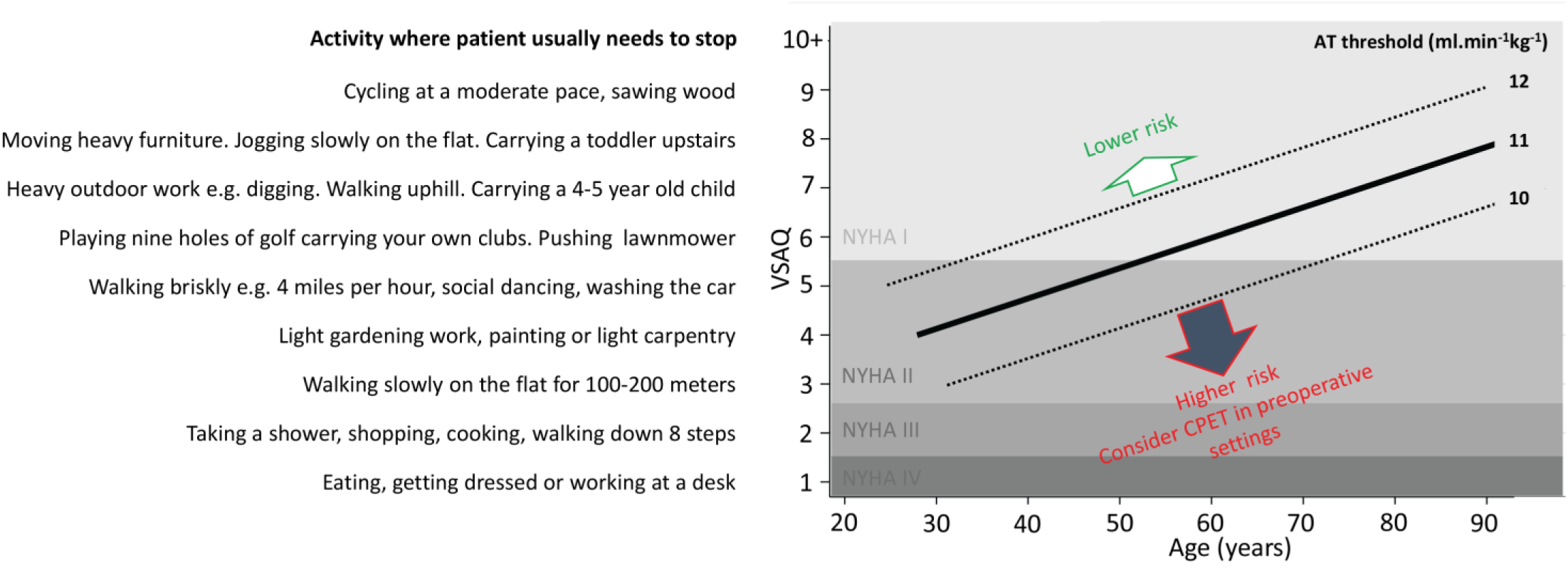
Age-VSAQ method suggesting lower and higher risk AT categories. *The lowest 10 VSAQ scores and associated exercise limitations are plotted against age. Horizontal bands indicate the respective New York Heart Association (NYHA[25]) categories of I (no symptoms on ordinary physical activity), II (limited on ordinary activity), III (limited at 20-100m), and IV (limited at rest). To indicate lower and higher risk categories, the regression line is plotted for an anaerobic threshold of 11 ml*.*min*^*- 1*^.*kg*^*-1*^. *To provide an indication of confidence limits, and the direction and scale of variation if this threshold scale were to be adjusted, regression lines for anaerobic thresholds of 10 and 12 ml*.*min*^*-1*^.*kg*^*-1*^ *are also plotted*

## Discussion

We demonstrate that high proportions of patients with a label that might be expected to mean “pre-existing cardio-respiratory condition” do not fall into classical high risk categories when more carefully evaluated. Furthermore, age and usual resting SpO_2_ were not associated with the anaerobic threshold and therefore cardiorespiratory risk status in continuous, or categorical analyses. However, lower haemoglobin, and haemoglobin-dependent indices of arterial oxygen content and delivery were important predictors of lower anaerobic threshold and a higher risk state: our *post hoc* calculations suggested increasing haemoglobin could have moved many of the high risk group into a lower risk category. We also demonstrate that a simple patient-based metric, the VSAQ, could allocate patients to lower and higher cardiorespiratory risk categories as based on the anaerobic threshold of <11 ml.min^-1^ kg^-1^.

The study numbers are small but notably demonstrated non-overlapping confidence intervals for key variables of oxygen tissue delivery. The current findings build on substantial previously published data and analyses on pulmonary AVM [16,17] and general population [5-10] cohorts. Furthermore, if the American Society of Anaesthetists (ASA) Physical Status Classification System [1] was employed, most pulmonary AVM patients would not fall into a high risk category because in the absence of other diseases, individuals with pulmonary AVMs rarely complain of respiratory symptoms.[14,17,22] Compensatory mechanisms are so effective that in one study, work rate and oxygen consumption on maximal CPET did not improve following embolisation treatments that obliterated the pulmonary AVM(s) and improved SpO_2_.[16] However, the issue is how best to capture this good exercise tolerance. We have previously reported the ease of use with the VSAQ in a pre-assessment clinic.[18] We have now adjusted to send the VSAQ to patients by email so they can report back the lowest number at which they needed to stop at a subsequent teleconsultation, thus conveying complex physiological information in seconds (*Onabanjo et al, manuscript in preparation*). While measurements in acute settings do not reflect the patient’s baseline, usual activity could be captured by the VSAQ either before or at the time of triage assessment.

Potential mechanisms for the association between lower anaerobic threshold and less successful surgical outcomes have been put forward, including the suggestion that regular exercise stimulates ischaemic preconditioning and lessens surgical demand by enabling the body to adjust to ischaemia and better utilise oxygen. Additionally, endurance exercise has been found to increase mitochondrial mass, which can therefore delay the start of anaerobic respiration by enhancing the utilisation of oxygen by mitochondria.[6] “Prehabilitation” or pre-operative exercise therapy has been found to improve post-operative outcomes in other disease groups and has been proposed to help prepare for COVID-19 infection.[23] Herein we also show that addressing anaemia is likely to be an additional strategy to reduce cardiorespiratory risk status.

In summary, high proportions of patients with a label that might be expected to mean “pre-existing cardiorespiratory condition” do not fall into classical high risk categories when more carefully evaluated. Given the need for appropriate allocation of ward/critical care resources, whether for surgery, or in infective setting, we suggest the VSAQ offers a cost-effective tool that can be easily integrated into triages or anaesthetic pre-assessments to assist with rapid evaluation of cardiorespiratory risk.

## Data Availability

Following peer-review publication, anonymised data will be available on request from the corresponding author.

## DECLARATIONS

### ETHICS APPROVAL AND CONSENT TO PARTICIPATE

-Ethical approvals in place:

1. CPET Studies: National Research Ethics Service (NRES) Committee South West London REC3: 11/H0803 and NRES London Riverside Committee:15/L0/0590, written informed consent for all
2. Case Notes review: Hammersmith and Queen Charlotte’s and Chelsea Research Ethics Committee (LREC 2000/5792, approved without requiring consent.

### CONSENT FOR PUBLICATION

- Not applicable

### AVAILABILITY OF DATA AND MATERIAL

- The datasets analysed during the current study are available from the corresponding authors on reasonable request

### COMPETING INTERESTS

- The authors have no competing interest to declare

### FUNDING

- Funding was received from the European Respiratory Society (2012 Rare Disease Achievement Award to CLS); National Institute of Health Research London (NW) Comprehensive Local Research Network; HHT patient donations; Imperial College BSc project funds (to CLS for ST, FG, TS, and VS), and the National Institute of Health Research Biomedical Research Centre Scheme (Imperial BRC). The views expressed are those of the authors and not necessarily those of funders, the NHS, the NIHR, or the Department of Health and Social Care.

### AUTHORS’ CONTRIBUTIONS

***-***Conception and design: ST, FG, TS, CLS. Analysis and interpretation: ST, FG, TS, JP, BM, JEJ, ST, VS, JM, HCT, LH and CLS. Drafting the manuscript for important intellectual content: ST, FG, CLS. *In detail:* ST devised the CPET analyses focussing on anaesthetic risk, performed literature studies, added to the pulmonary AVM CPET database, performed the data analysis, generated Figures 2, 3 and Supplementary Figures 1 and 2, and wrote the first draft of the manuscript. FG introduced the VSAQ clinic assessments, performed literature studies, generated the observational database, drafted manuscript sections and generated Supplementary Figure 3. TS performed literature studies, assisted in obtaining ethical approvals to recruit patients with airflow obstruction, and added to the pulmonary AVM CPET database. JP co-supervised ST, and FG, and performed and interpreted CPET data measurements. VS performed literature studies, contributed to generation of the observational pulmonary AVM database, assisted in obtaining initial ethical approvals to recruit patients without airflow obstruction, and initiated the pulmonary AVM CPET database. JM generated the VSAQ, advised on physiological concepts, and contributed to data interpretation. HT co-supervised ST, and FG, and performed and interpreted CPET data measurements. LH co-supervised TS and VS, and performed CPET data measurements. CLS supervised all students, devised the initial CPET study, reviewed all patients, performed literature searches, analysed and interpreted data, performed additional presented data analyses, generated other Figures, and wrote the final manuscript. All authors contributed to and approved the final version of this manuscript.

## ACKNOWLEDGEMENTS

**-**The authors thank the patients for their willing cooperation in these studies.

## Supplementary Data

### Supplementary Methods

#### METS VSAQ AT Formulae

*Since*

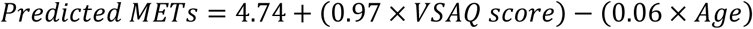

*And*

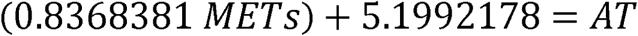

*Then*

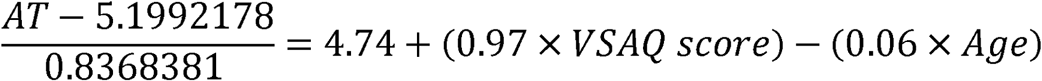

*And for the AT cut off of 11ml*.*min*^*-1*^*kg*^*-1*^

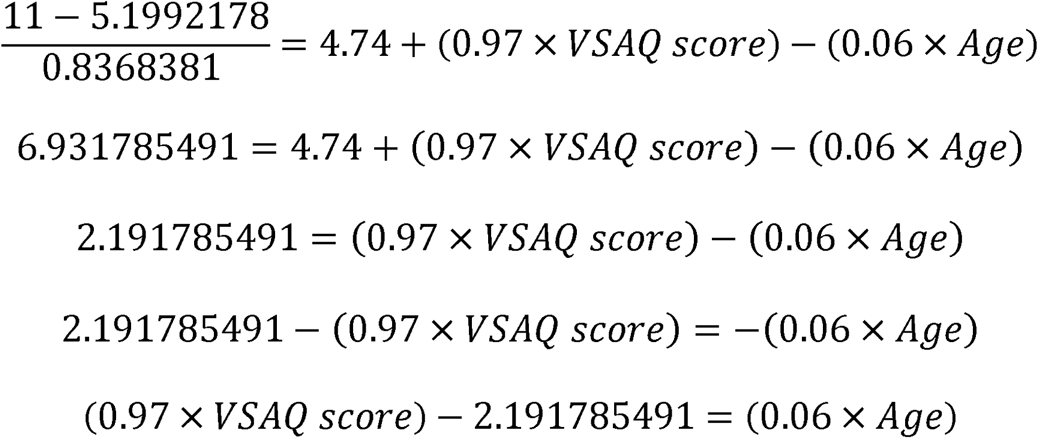

*So by Excel, Tested ages where the relevant VSAQ score would yield a figure <2*.*19:*

| VSAQ | VSAQx.97 | (VSAQx.97)-2.191785491 | (VSAQx.97)-2.191785491/0.06 |
| --- | --- | --- | --- |
| 1 | 0.97 | -1.221785491 | -20.3631 |
| 2 | 1.94 | -0.251785491 | -4.19642 |
| 3 | 2.91 | 0.718214509 | 11.97024 |
| 4 | 3.88 | 1.688214509 | 28.13691 |
| 5 | 4.85 | 2.658214509 | 44.30358 |
| 6 | 5.82 | 3.628214509 | 60.47024 |
| 7 | 6.79 | 4.598214509 | 76.63691 |
| 8 | 7.76 | 5.568214509 | 92.80358 |
| 9 | 8.73 | 6.538214509 | 108.9702 |
| 10 | 9.7 | 7.508214509 | 125.1369 |
| 11 | 10.67 | 8.478214509 | 141.3036 |
| 12 | 11.64 | 9.448214509 | 157.4702 |
| 13 | 12.61 | 10.41821451 | 173.6369 |

**Supplementary Figure 1.**
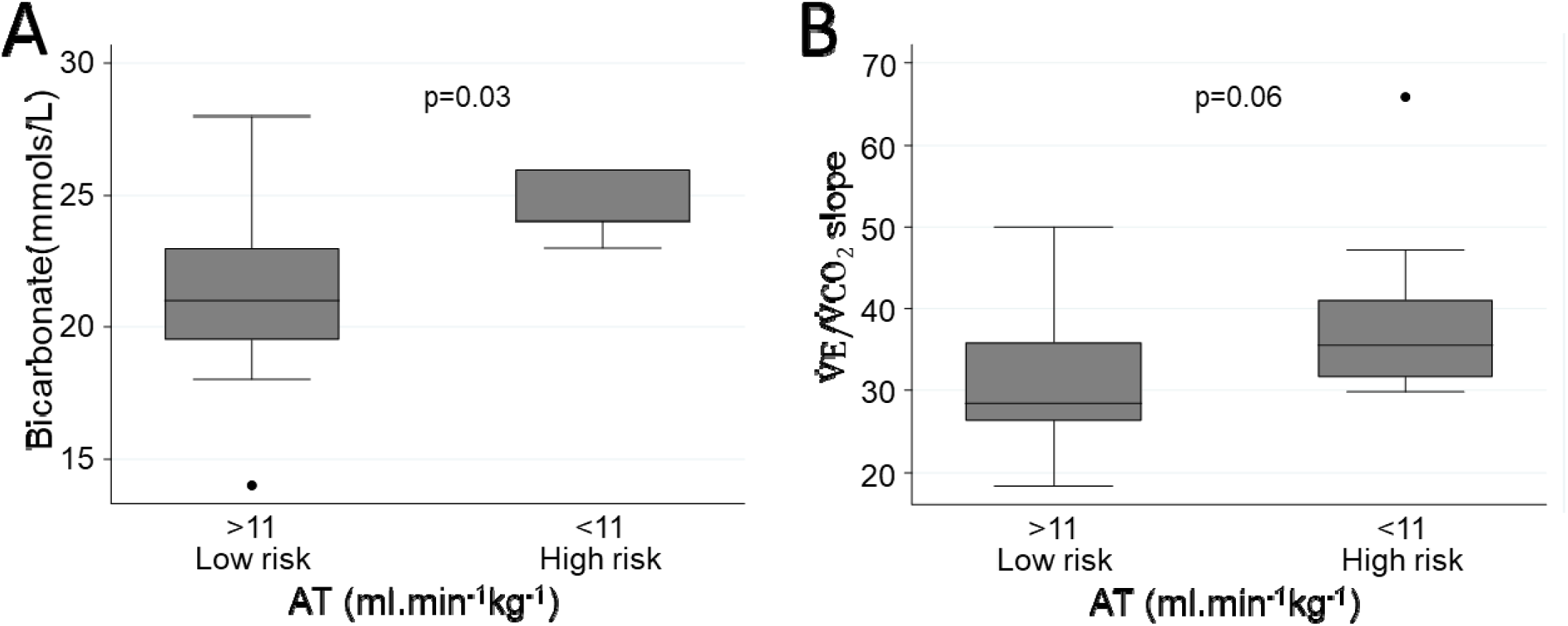
Comparison of CPET-derived variables related to ventilation in low and high risk groups delimited by anaerobic threshold (AT) of 11ml.min^-1^kg^-1^: **A)** serum bicarbonate (mmol/L) available in N=17, **B)** V_E_/V_CO2_ slope. Boxes indicate the median and interquartile range (IQR), and error bars represent 2 standard deviations, with dots at the extremes representing outliers. P-values were calculated by Mann Whitney *U* test. Note that the lower risk group displayed lower bicarbonate indicative of more exuberant ventilation, yet had less steep V_E_/V_CO2_ slope consistent with greater ventilatory efficiency.

**Supplementary Figure 2.**
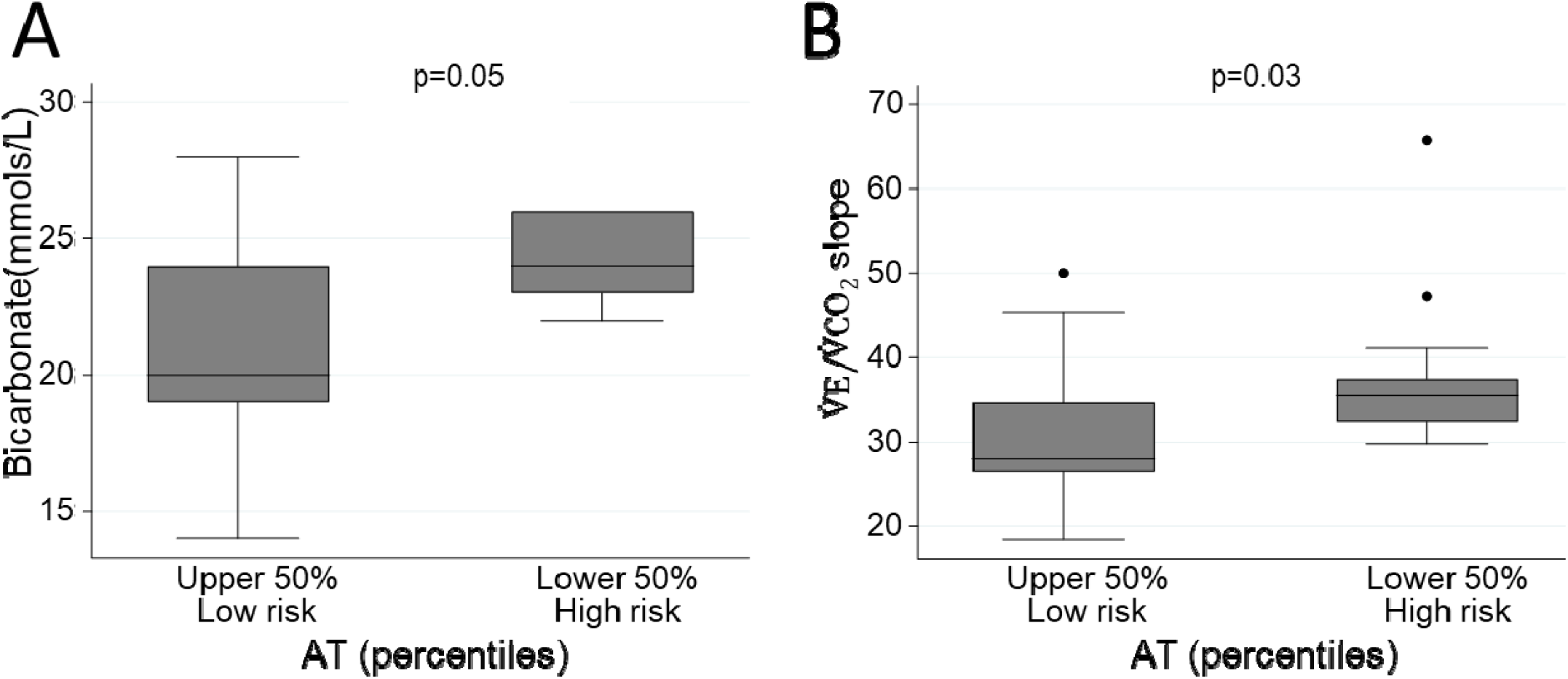
Comparison of CPET-derived variables related to ventilation in low and high risk groups delimited by above and below median anaerobic threshold (AT): **A)** Serum bicarbonate (mmol/L) available in N=17, **B)** V_E_/V_CO2_ slope. Additional details as for Supplementary Figure 1 above.

**Supplementary Figure 3.**
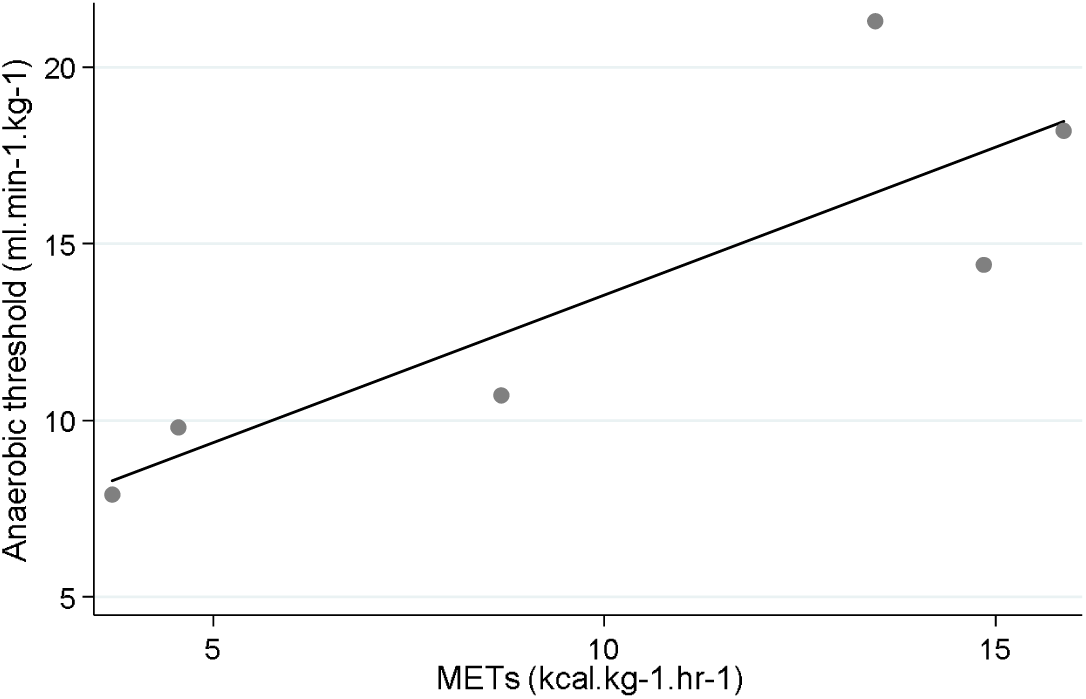
Relationship between METs and Anaerobic Threshold. Anaerobic threshold plotted against predicted METs from VSAQ score. The regression coefficient was 0.84 (95% confidence intervals 0.12, 1.56), p= 0.032.

## Notes

### Competing Interest Statement

The authors have declared no competing interest.

## REFERENCES

1 National Institute for Health and Care Excellence Routine preoperative tests for elective surgery Available from: https://wwwniceorguk/guidance/ng45

2 Agnew N. Preoperative cardiopulmonary exercise testing. Continuing Education in Anaesthesia Critical Care and Pain. 2010; 10(2): 33-37. Available from: doi: 10.1093/bjaceaccp/mkq001.

3 Ridgway ZA, Howell SJ Cardiopulmonary exercise testing: a review of methods and applications in surgical patients European Journal of Anaesthesiology 2010;27(10): 858–865 Available from: doi:101097/EJA0b013e32833c5b05

4 Moran J, Wilson F, Guinan E, Mccormick P, Hussey J, Moriarty J Role of cardiopulmonary exercise testing as a risk-assessment method in patients undergoing intra-abdominal surgery: a systematic review British Journal of Anaesthesia 2016;116(2):177–191 Available from: doi:101093/bja/aev454

5 Myers J, Do D, Herbert W, Ribisl P, Froelicher VF. A nomogram to predict exercise capacity from a specific activity questionnaire and clinical data. Am J Cardiol. 1994;73(8): 591–596.

6 Myers J, Bader D, Madhavan R, Froelicher V. Validation of a specific activity questionnaire to estimate exercise tolerance in patients referred for exercise testing. Am Heart J 2001;142(6): 1041–1046.

7 Athanasopoulos LV, Dritsas A, Doll HA, Cokkinos DV.The role of NT-proBNP in explaining the variance in anaerobic threshold and VE/VCO(2) slope. J Cardiopulm Rehabil Prev. 2011 Sep-Oct;31(5):316–21.

8 Jette M, Sidney K, Blümchen G. Metabolic equivalents (METS) in exercise testing, exercise prescription, and evaluation of functional capacity. Clin Cardiol. 1990;13(8): 555–565.

9 McAuley P, Myers J, Abella J, Froelicher V. Evaluation of a specific activity questionnaire to predict mortality in men referred for exercise testing. Am Heart J. 2006;151(4): 890. e1-890. e7.

10 Snowden CP, Prentis JM, Anderson HL, Roberts DR, Randles D, Renton M, Manas DM. Submaximal cardiopulmonary exercise testing predicts complications and hospital length of stay in patients undergoing major elective surgery. Ann Surg. 2010 Mar;251(3):535–41.

11 Shovlin CL, Condliffe R, Donaldson JW, Kiely DG, Wort SJ; British Thoracic Society British Thoracic Society Clinical Statement on Pulmonary Arteriovenous Malformations Thorax 2017 Dec;72(12):1154–1163

12 Shovlin CL. Pulmonary arteriovenous malformations. Am J Respir Crit Care Med. 2014 Dec 1;190(11):1217–28.

13 Whyte MK, Hughes JM, Jackson JE, Peters AM, Hempleman SC, Moore DP, et al. Cardiopulmonary response to exercise in patients with intrapulmonary vascular shunts. J Appl Physiol 1993;75:321–328

14 Santhirapala V, Williams LC, Tighe HC, Jackson JE, Shovlin CL. Arterial oxygen content is precisely maintained by graded erythrocytotic responses in settings of high/normal serum iron levels, and predicts exercise capacity. An observational study of hypoxaemic patients with pulmonary arteriovenous malformations. PLoS ONE 2014 Mar 17;9(3):e90777.

15 Rizvi A, Babawale L, Tighe HC, Macedo P, Hughes JMB, Jackson JE, Shovlin CL. Hemoglobin is a vital determinant of arterial oxygen content in hypoxemic patients with pulmonary arteriovenous malformations. Ann Am Thorac Soc. 2017 Jun;14(6):903–91.

16 Howard LSGE, Santhirapala V, Murphy K, Mukherjee B, Busbridge M, Tighe HC, et al. Cardiopulmonary exercise testing demonstrates maintenance of exercise capacity in patients with hypoxemia and pulmonary arteriovenous malformations. Chest. 2014;146(3): 709–718. Available from: doi:10.1378/chest.13-2988.

17 Gawecki F, Strangeways T, Amin A, Perks J, Wolfenden H, Thurainatnam S, Rizvi A, Jackson JE, Santhirapala V, Myers J, Brown J, Howard LSGE, Tighe HC, Shovlin CL. Exercise capacity reflects airflow limitation rather than hypoxaemia in patients with pulmonary arteriovenous malformations. QJM. 2019 Jan 17. doi: 10.1093/qjmed/hcz023.

18 Gawecki F, Myers J., Shovlin CL. Veterans Specific Activity Questionnaire (VSAQ): a new and efficient method of assessing exercise capacity in patients with pulmonary arteriovenous malformations BMJ Open Respir Res. 2019 Mar 1;6(1): e000351 http://bmjopenrespres.bmj.com/cgi/content/abstract/6/1/e000351

19 Vorselaars VM, Velthuis S, Mager JJ, Snijder RJ, Bos WJ, Vos JA, et al. Direct haemodynamic effects of pulmonary arteriovenous malformation embolisation. Neth Heart J 2014;22:328–333.

20 Shovlin CL, Buscarini E, Kjeldsen AD, Mager HJ, Sabba C, Droege F, Geisthoff U, Ugolini S, Dupuis-Girod S. European Reference Network For Rare Vascular Diseases (VASCERN) Outcome Measures For Hereditary Haemorrhagic Telangiectasia (HHT). Orphanet J Rare Dis. 2018 Aug 15;13(1):136. doi: 10.1186/s13023-018-0850-2.

21 Finnamore H, Le Couteur J, Hickson M, Busbridge M, Whelan K, Shovlin CL. Hemorrhage- adjusted iron requirements, hematinics and hepcidin define hereditary hemorrhagic telangiectasia as a model of hemorrhagic iron deficiency. PLoS One. 2013 Oct 16;8(10):e76516.

22 Santhirapala V, Chamali B, McKernan H, Tighe HC, Williams LC, Springett JT, Bellenberg HR, Whitaker AJ, Shovlin CL. Orthodeoxia and postural orthostatic tachycardia in patients with pulmonary arteriovenous malformations: a prospective 8-year series. Thorax. 2014 Nov;69(11):1046–7.

23 Shovlin CL, Moorthy K, Lees C. Covid-19: Home based exercise activities could help during self isolation BMJ 2020: https://blogs.bmj.com/bmj/2020/03/16/covid-19-home-based-exercise-activities-could-help-during-self-isolation/

24 Thurainatnam S. Pre-Operative Insights from Cardiopulmonary Exercise Testing in Patients with Pulmonary Arteriovenous Malformations. BSc Project, Imperial College London (2017).

25 Criteria Committee, New York Heart Association, Inc. Diseases of the Heart and Blood Vessels.Nomenclature and Criteria for diagnosis, 6th edition Boston, Little, Brown and Co. 1964, p 114.

